# Real-Time Biometric Monitoring for Cognitive Workload Detection: A Narrative Review of Applications in High-Demand Professions

**DOI:** 10.1101/2025.08.28.25334668

**Authors:** Reginald B. O’Hara, Shelby Loftis, Cynthia Rando

## Abstract

This narrative review examines the theoretical foundations of mental workload, evaluates biometric monitoring methods, addresses ethical and privacy issues, and highlights future directions for longitudinal research. We focus on the integration of wearable sensors, multimodal data, artificial intelligence (AI), and machine learning (ML) frameworks to enhance adaptive task scheduling and safety in cognitively demanding professions.

Continuous, real-time monitoring through wearable devices and multidimensional data analysis show promise for identifying and managing cognitive overload before it degrades performance. Despite this potential, significant challenges exist, including data protection, sensor reliability, calibration consistency, information processing, network capabilities, and variability in individuals’ responses. Physiological and behavioral measures as well as subjective and performance indicators offer valuable insights into the early signs of cognitive strain, suggesting that biometric monitoring could help organizations detect performance decline sooner.

Evidence shows that these technologies are feasible in professions that require high precision, rapid decision-making, and sustained attention. However, only sparse longitudinal comparisons exist regarding the effectiveness of different biometric tools in real-world operational contexts, particularly with respect to data security and standardization.

Integrating physiological and behavioral data with subjective assessments analyzed through AI and ML may enable early warning signs for overload in both individuals and teams working in high-stress, time-critical settings. Such approaches could inform work-recovery cycles, reduce error rates, and sustain cognitive performance. Further empirical research is necessary to confirm sensor accuracy in applied environments and to validate AI and ML predictions before large-scale deployment in sectors such as air traffic control, public safety, healthcare, and industrial operations.

**Graphical Abstract:** 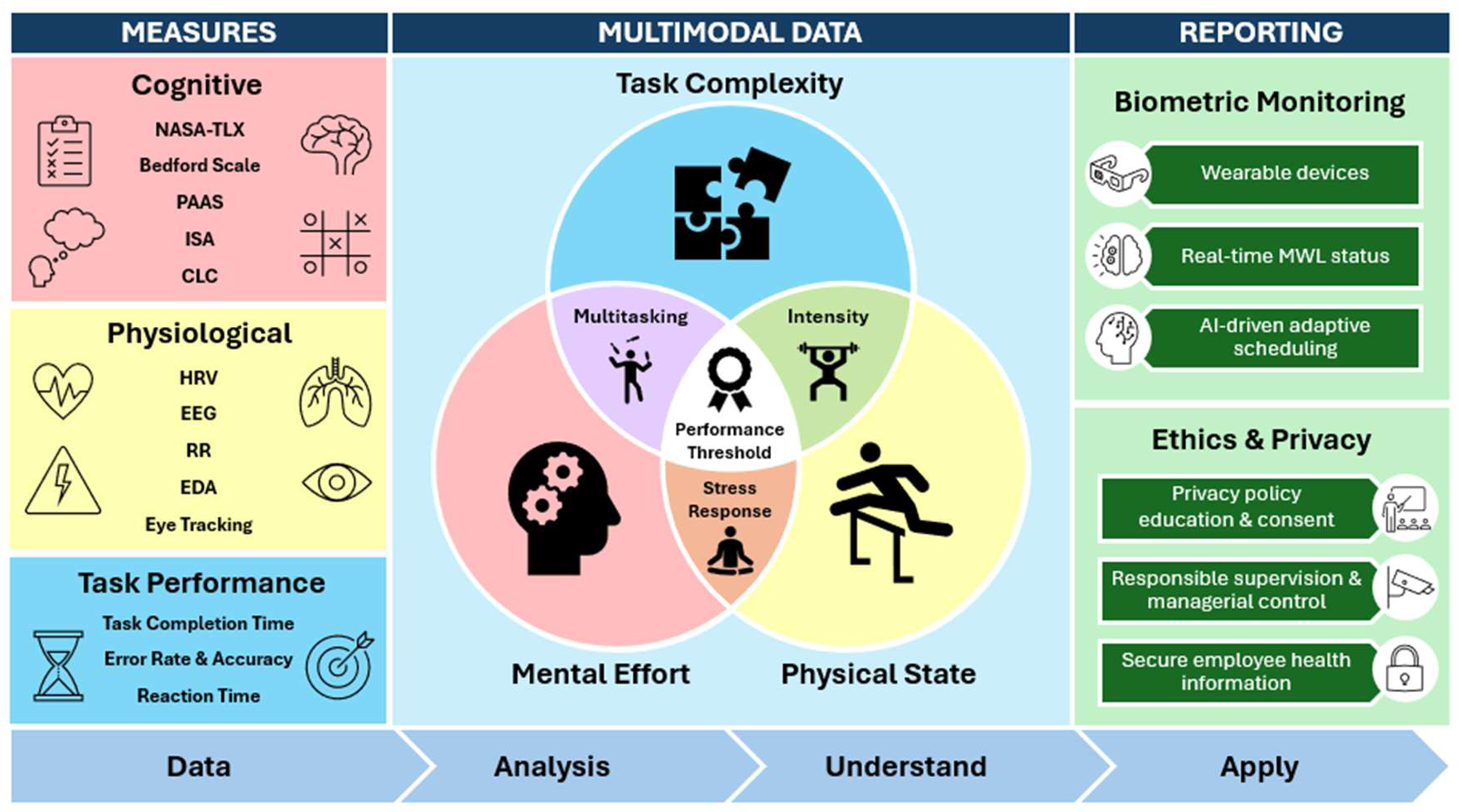

## 1. Introduction

Mental workload (MWL), often termed cognitive workload, refers to the cognitive effort required to effectively perform and complete a task at a specific moment [18]. MWL is influenced by task complexity, mental effort, and time pressure to meet task goals [18,20,32]. The human brain works similar to a computer; when overwhelmed by excessive or competing inputs, such as during multitasking, it becomes prone to errors and slower execution.

In recent years, employee health and stress have increased markedly, likely because of rapid technological growth and advancements in information technology (IT), such as artificial intelligence (AI), robotic process automation, remote work platforms, team collaboration tools, and workforce analytics, which continuously track employee performance [20]. Although these evolving technologies are helpful, they can also increase employees’ cognitive demands. For example, the increase in clinical settings and the integration of IT, including electronic medical records, patient monitoring systems, and communication tools, is designed to improve the delivery of patient care. However, these systems require clinicians and nurses to divide their attention between multiple platforms and patient-care tasks, thereby encouraging multitasking.

Frequent task-switching can overload mental resources, leading to increased error rates [11]. Multitasking increases the mental load by pushing the brain beyond its cognitive reserves, often resulting in task errors, safety risks, and cognitive processing delays, particularly in high-pressure ecosystems where rapid and accurate performance is required. Overload likely occurs not because a person can truly focus on multiple tasks at once but because multitasking forces the brain into immediate task switching, which depletes brain resources, derails attention, and increases the risk of task mistakes [18].

The emerging interest and increasing adoption of wearable biometric sensors play a critical role in enabling rapid collection, processing, and analysis of neurophysiological and behavioral pattern data in real-time (RT) contexts [11,20,18]. As mental workload increases, so does the risk of human error, especially in work settings that require sustained attention and rapid decision making under time constraints. To counteract these risks, non-invasive biometric monitoring systems offer a promising solution by enabling rapid responses and individualized targeted interventions. These systems assess the RT indicators of brain activity, heart rate, heart rate variability (HRV), pupil dilation, and fixation to detect early signs of overload [7,11,18,39]. By continuously monitoring these physiological signals, biometric sensors can help mitigate critical errors, improve employee safety and health, and sustain productivity, thereby enhancing the performance metrics.

Individuals in demanding roles can greatly benefit from RT biometric monitoring systems, which help prevent injuries, sustain productivity, preserve health, and minimize errors. For instance, construction workers often experience significant occupational fatigue because of the mentally and physically demanding nature of their work, which often requires working in many different types of environments (e.g., outside) [29]. Fatigue leads to a higher risk of errors and diminished awareness of potentially hazardous situations, resulting in catastrophic consequences.

In 2019 alone, over 200,000 injuries and illness cases were reported among construction workers, along with 79,700 missed workdays, all of which negatively affected productivity [29]. These findings underscore the importance of addressing occupational fatigue not only in construction but also in other demanding occupations such as nursing, emergency responders (fire, police, EMS), and specialized industrial technicians [13,15,29]

Integrating multimodal sensor datasets with fatigue measurement systems to capture RT biometric data can play a critical role in the early detection of mental fatigue [7,39]. This proactive approach helps to prevent errors, supports productivity, and sustains both physical and cognitive health. However, in many high-pressure occupations, employees often lack access to RT monitoring systems that can detect mental overload before it leads to task errors, declining mental health, or decreased performance [1]. Mental and physical stress data are often collected retrospectively, only after a series of stressful events, and typically rely on subjective mental task rating scales without the inclusion of objective RT biometric data for evaluation [18].

The central aim of this review was to assess the scientific, technological, and practical inclusion of RT biometric monitoring systems for the early detection and management of MWL in high-demand occupations. The objectives of this review are to: 1) classify the theoretical underpinnings of MWL; 2) assess various biometric modalities and performance indicators; 3) demonstrate the potential application of AI and machine learning (ML) to further enhance RT monitoring accuracy; 4) address ethical, security, and privacy challenges; and 5) suggest future empirical research directions.

### 1.2. Statement of significance

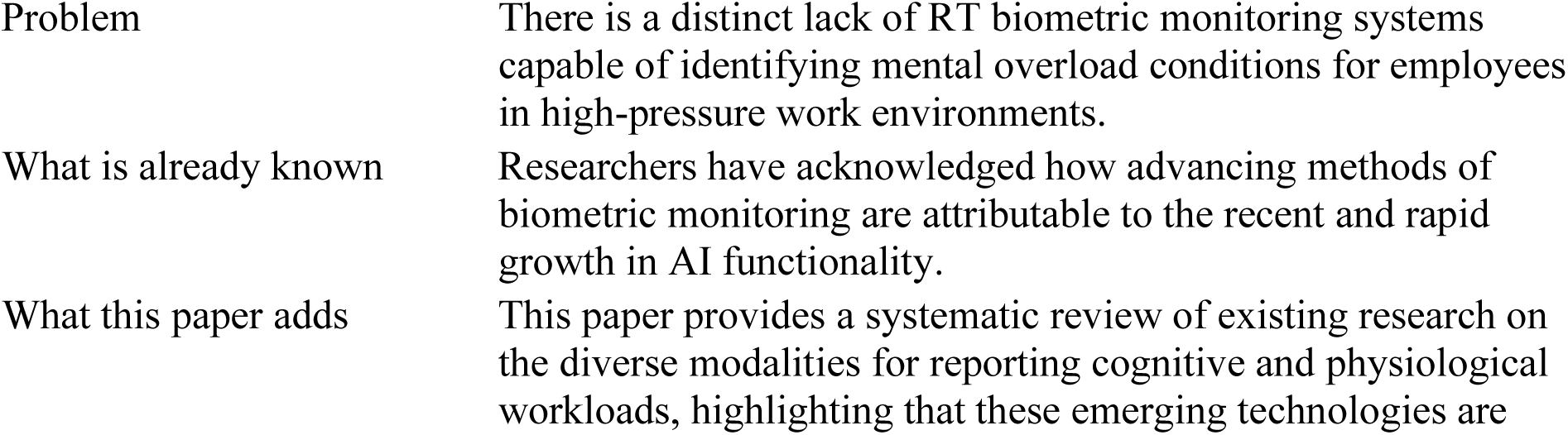
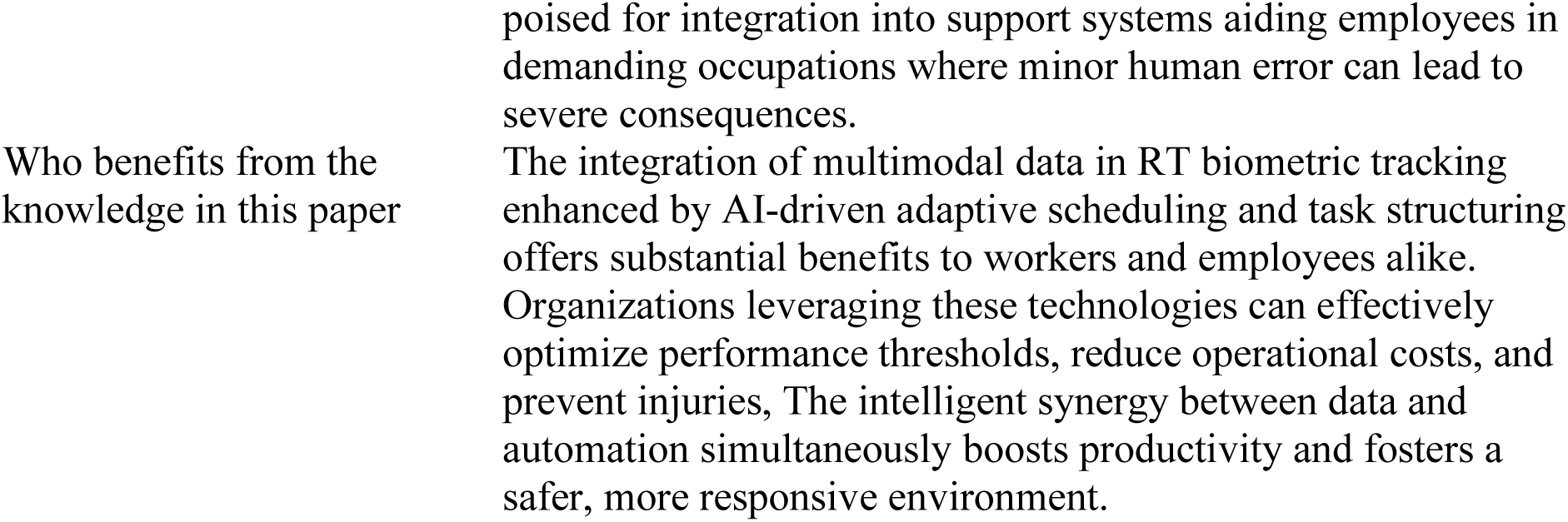

## 2. Methods

### 2.1 Search Strategy

We structured this Narrative Review (NR) according to Ferrari’s [12] guidance on methodological rigor in non-systematic reviews alongside NR quality criteria developed by Baethge et al. [4]. The literature was sourced from Google Scholar, PubMed Central, and electronic university databases between 1981 and 2025, focusing on open-access peer-reviewed articles relevant to cognitive/mental workload, biometric monitoring, and multimodal data. Our aim was to locate relevant, diverse sources aligned with the central aim and objectives of this review to evaluate real-time biometric monitoring in high-demand work environments.

### 2.2. Inclusion Criteria for Selection of Studies

The criteria for selecting the sources were established and aligned with the central aim and objectives of this review, as shown in **Table 1**. Only open-access, full-text, peer-reviewed articles were considered. This review was conducted using a university electronic library (MINERQUEST) and PubMed Central database, which provided unrestricted open access to the library catalog and database using keyword searches. We performed two searches on Google Scholar and PubMed Central, which aided in identifying the initial and final records. Additional records were sourced based on our inclusion criteria (full-text availability) relevant to biometric monitoring systems, mental/cognitive workload, mental stress, and multiple modalities using AI or ML, and studies focused on using multimodal data relevant to cognitive workload.

**Table 1.** Inclusion criteria.

| Inclusion Criteria |
| --- |
| 1. Research articles |
| 2. Systematic reviews |
| 3. Defense technical reports |
| 4. Free full-text, open access peer reviewed articles |
| 5. Populations(s): high demand occupations, robotic tech and smart factory |
| 6. Context: use of multimodal data, use of biosensor modalities in workplace |
| 7. Integration of biometric sensors for real-time monitoring of mental workload |
| 8. Published between 1981-2025 |
| 9. Written in the English language |
| 10. Not published as a conference abstract or poster presentation |

### 2.3. Search Strategy and Data Extraction

A primary literature search on Google Scholar using Boolean operators to separate the search terms “biometric sensors” or “cognitive/mental” and “multimodal data” load biometric sensors” from 1981 to 2025, focusing on review articles, systematic reviews, observational studies, and government technical reports from free full-text, open-access articles yielded 39 selected results.

We determined the key concepts for the review and navigated databases for appropriate keywords directly related to the topics of interest [12]. Zotero software was used to extract the results for the initial search strategy in the Google Scholar advanced search filter by entering the search terms “biometric sensors”, which yielded 10,200 results. The second search focused on “cognitive workload” AND “multimodal data,” generating 755 combined results. For the third search, we applied the key terms “mental workload” and “occupational stress,” which produced 3,422 results. A fourth Google Scholar advanced search highlighted the terms “artificial intelligence,” “predictive analytics,” AND “electronic health records,” yielding 16,800 articles.

The same search strategy was performed in the electronic university library in the MINERQUEST search bar, choosing PubMed Central as the secondary database search, using the terms “biometric sensors” OR “cognitive workload” AND “multimodal data” for the period 1981–2025. The first search yielded 9,230 results with varying degrees of relevance to specific inclusion criteria. We applied filters for the second search using only the available online and peer-reviewed data. We narrowed the resource type to only articles, review articles, reports, and first online articles, which yielded 946 results. The first two authors (RBO and SL) independently searched for primary and secondary sources, screened titles and abstracts, and assessed the full text of published articles potentially relevant to the narrative review topic and objectives.

**Fig. 1.**
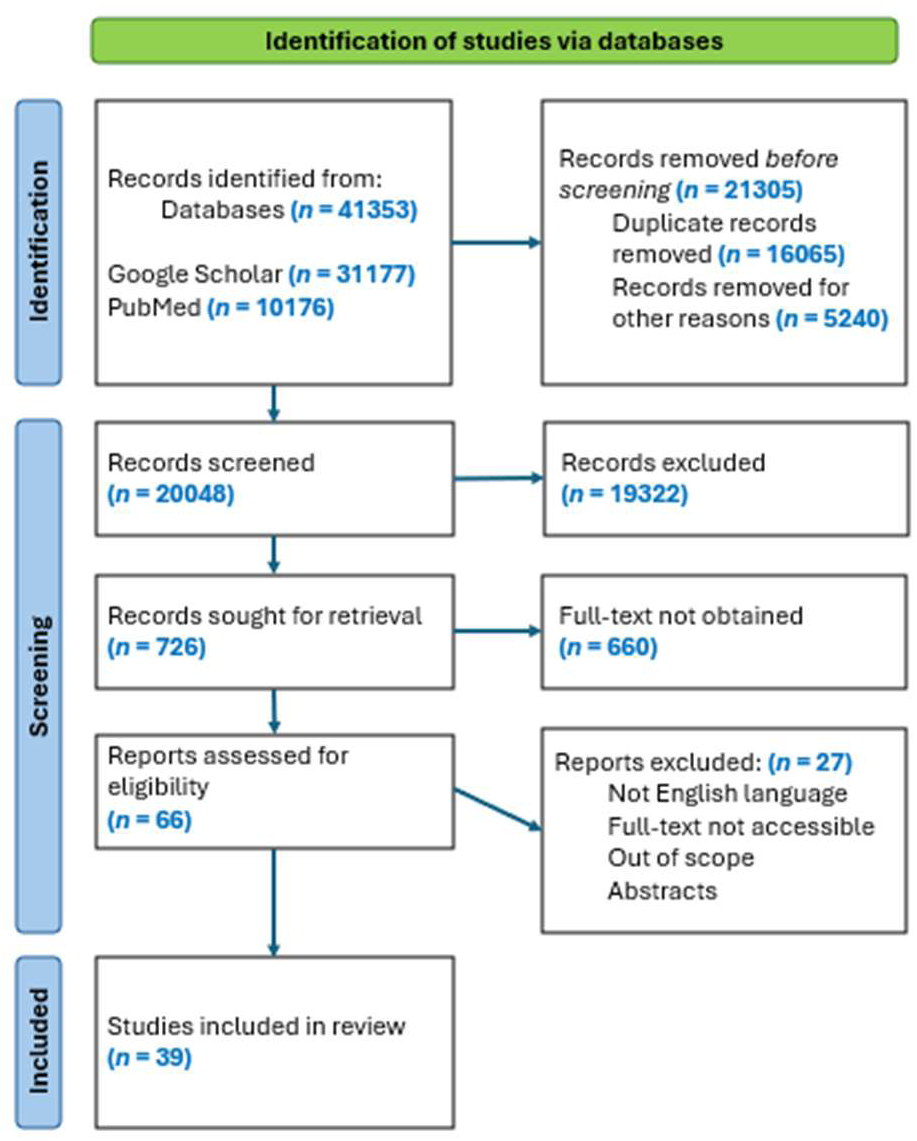
PRISMA Flow Diagram [25].

We then applied filters to narrow the subject to only 1) technology, 2) physical sciences, 3) machine learning, 4) physiology, 5) artificial intelligence, 6) human beings, and 7) methods, yielding 762 results. We then refined the results to include only open access and narrowed the subject search to science, research, review, and data collection, which yielded 66 results. Zotero software was used to extract the results. The 66 results from PubMed Central and Google Scholar were compared to remove all remaining duplications and borderline studies, which were assessed according to the inclusion criteria and specifically to population (high-demand occupations) and were not occupationally focused or lacked inadequate biometric validation studies.

## 3. Results

All the articles were assessed for eligibility based on the inclusion criteria listed in **Table 1**. Of the examined articles, 39 were selected for inclusion. The sources selected for inclusion were review articles (n = 19), primary research articles (n = 18), systematic reviews (n = 2), defense technical reports (n = 2), and a doctoral thesis (n = 1).

## 4. Discussion

### 4.1. Theoretical Foundations

The theoretical underpinnings supporting mental (cognitive) overload originate from the work of Wickens and Sweller, derived from multiple resource theory (MRT) and cognitive load theory (CLT) [37,35]. For example, Wickens [37] showed that the brain allots separate resources, such as auditory and visual, to different types of tasks, which can result in task interference when those tasks require cognitive resources from multiple brain channels. This explains why multitasking can be effective on some occasions but not on others. Rather than viewing mental resources as a single fuel reserve, MRT suggests that human information processing is divided across cognitive channels [37]. This segmentation helps elucidate why multitasking may succeed in some situations and fail in others.

Wickens [37] proposed that the human brain has separate channels for executing different task types. For instance, driving requires visual-spatial resources such as watching the road and steering, whereas talking uses auditory-verbal resources such as listening and speaking. Although humans can typically manage tasks that engage in varying sensory modalities (auditory and visual), performance depends not only on the type of channel, but also on situational awareness and self-regulation. Research has shown that when conversing with a passenger, people routinely engage in a form of mutual self-monitoring, whereby passengers may pause the conversation during complex driving maneuvers, resulting in a reduced cognitive load [14]. However, during cell phone conversations, remote individuals lack visual access to the driving environment, which prevents adaptive regulation of conversation [14]. This absence of situational awareness often contributes to greater distraction and performance decline, even when different modalities are used [14,27,37].

Sweller [35] advanced Wickens’ work through CLT, which showed that a person’s brain has a limited capacity to learn new things simultaneously, especially when dealing with complex information. He identified three types of mental (brain) loads relevant to CLT: 1) intrinsic load, which is the inherent difficulty of what an individual is attempting to learn; 2) extraneous load, which is the unnecessary stuff that gets in the way, such as poor instructions or poor design of dashboards or protocols; and 3) germane load, which is the mental effort required for a person to build understanding or develop mental models to organize steps in the mind or connect new ideas with old ones [35].

Employers who use CLT to improve employee performance may consider schema-based training, whereby workers are trained with realistic, repeated scenarios, allowing them to build automatic responses and reduce the intrinsic load during a real crisis [24]. Simplifying interfaces and tools can reduce extraneous load by designing clear dashboards or protocols that eliminate the need to guess or decode complex information under pressure [27]. .Large information dumps can decrease brain load when broken down into step-by-step flows. The use of external memory aids, such as visual cues, reminders, or auto-calculations, can also help ease the pressure on the working memory [24].

### 4.2. Biometric Sensor Modalities & Mental Workload

This section describes the commonly used evaluation measures and biometric sensor modalities derived from scientific literature for the continuous monitoring of MWL in RT and real-world work environments. The introduced biometric indicators are not all-inclusive, as they can vary depending on the type, complexity, or demand of the required occupational task. The overarching goal was to collect reliable data that showed only minor changes from one day to the next. The devices used to gather this data must also be valid and accurately measure what they are intended to measure.

As MWL is a complex concept that differs across professions, it is insufficient to measure the amount of work done by someone. Consideration should also be given to the mental demands, stress, and complexity of the job tasks. To enhance predictability and capture a more comprehensive and accurate picture of MWL, it is necessary to integrate diverse types of data and biometric sensing modalities such as physiological signals (e.g., heart rate variability), performance results, self-reported experiences, and task difficulty. Therefore, the evaluation measures discussed in each section should be viewed as essential parts of a larger MWL.

### 4.3. Physiological & Behavioral Measures

#### Heart Rate Variability (HRV)

HRV is a useful method for measuring sensitivity to physical and mental stressors. HRV measures the functioning of the sympathetic and parasympathetic branches, which in turn reflect the overall functionality of the autonomic nervous system (ANS) when an individual encounters a stressor [10,19]. Monitoring HRV provides insights into whether SNS and PNS branches function in a balanced manner. A high MWL can disrupt the balance between the two branches when the time and complexity of a mental task exceeds the brain reserves. Although HRV allows for RT monitoring of MWL, the repeatability of this measure can be affected by respiration rate, sleep rhythms, overall physical fitness level, and emotional state, all of which are considered confounding factors and can produce inconsistencies in work environments where physical exertion and stress levels may often change [18].

#### Electroencephalogram (EEG)

EEG measures electrophysiological signals through electrodes, often embedded into a cap and placed on the scalp, capturing RT electrical brain activity by detecting changes in brain wave patterns that correlate with mental effort, stress, and fatigue [5,18]. EEG is routinely used to objectively assess cognitive workload during tasks, providing more frequent tracking of time-dependent changes in brain activity without affecting actual performance [5,36].

#### Respiratory Rate (RR)

RR patterns depict the relationship between workload and stress by capturing the frequency and pattern of breathing, which change with mental or emotional stress. An increased workload typically elevates the breathing rate and reduces variability. By analyzing these deviations, respiratory monitoring offers a non-invasive physiological marker of stress and cognitive demand during task execution [20].

#### Electrodermal Activity (EDA)

EDA can signal the early detection of stress and emotional surges by assessing alterations in skin conductance resulting from the stimulation of sweat glands, which are regulated by the body’s stress stress-response system [7,18]. Elevations in EDA are indicative of increased stress, MWL, and emotional arousal. EDA is a non-intrusive, wearable technology for collecting unconscious physiological responses to cognitive demands in high-stake settings [7].

#### Eye Tracking

Even less invasive assessment indicators, such as eye blink and fixation rates, can be checked using eye-tracking monitors, which examine how long a person looks, including blink rate, pupil diameter, and fixation time [18]. With increased MWL, fixation patterns become more focused, and the blink rate decreases. This measure offers insight into attention, visual processing, and cognitive load during the execution of complex tasks or the use of a worker interface [18].

### 4.4. Subjective and Performance Measures

#### NASA-TLX (Task Load Index)

This multidimensional assessment tool evaluates the perceived physical and mental workload during a task, especially in critical environments such as spaceflight, aviation, healthcare fields, first responders, and human-computer interactions [16]. It can also be used in everyday work settings to identify tasks that are too demanding or frustrating. The questionnaire asks participants to subjectively rate six dimensions such as mental and physical demand, temporal demand, effort, and level of frustration [27].

After completing a specific task, the individual must provide a personal rating using six dimensions from 0 (very low) to 100 (very high), and a total score is calculated to assess the person’s overall workload [20]. The higher the number, such as a score closer to 100, the greater the MWL [18].

#### Bedford Scale

The Bedford Workload Scale is a 10-point tool originally developed to assess the mental workload of military, civil, and airline pilots during and directly after demanding flight tasks [28]. A score of 1 indicates a minimal workload, whereas a score of 10 indicates an excessive workload that prevents task completion. This simple scale design makes it practically useful in time-sensitive, high-performance environments such as aviation, where continuous task execution is critical [28].

#### Physical Activity Affect Scale (PAAS)

PAAS measures emotional responses to physical activity, helping to examine how physical activity or fatigue affects mood and, in turn, alters cognitive performance [18]. PAAS assesses emotional responses to physical activity, including feelings of energy, fatigue, tension, and relaxation. This is beneficial for jobs that require both physical exertion and mental tasks, such as first responders, industrial factory workers, and medical professionals [18].

#### Instantaneous Self-Assessment (ISA)

The ISA was originally developed for the aviation sector and captures self-assessments of perceived cognitive load in real-time [18]. It measures mental effort, demand, and stress during ongoing tasks and provides a simple yet effective measure of mental strain without disrupting task performance [18].

#### Cognitive Load Component Questionnaire (CLC)

The CLC questionnaire divides MWL into three key components: 1) mental effort, 2) task complexity, and 3) time pressure [31]. CLC reveals which specific cognitive demands are most demanding during task execution, offering valuable insights into how varying types of cognitive strain contribute to overall workload perception [31].

#### Task Completion Time

The influence of MWL on completion time can help determine how quickly an individual completes a task at varying MWL levels. An increased workload typically slows down performance, indicating cognitive strain or divided attention [18,32]. Assessing the task completion time is useful for evaluating the efficiency and adaptability of individuals in task execution within their specific job roles.

#### Error Rate and Accuracy

The error rate and accuracy measure the quality of task performance. A high MWL often leads to more errors and lower precision [18]. Monitoring these outcomes provides insights into how cognitive demands affect attention, decision-making, and task reliability, particularly in high-stake or detail-oriented work environments [1,7].

#### Reaction Time

Reaction time refers to the speed at which a person responds to a stimulus, such as a signal or sudden event. When the brain is under greater mental pressure, people usually react more slowly to the stimuli. This delay occurs because their thinking is overloaded. Reaction time is crucial for jobs that require rapid and precise responses [7,18].

### 4.5. Multisource Data Integration Methods

Multimodal assessment methods (MAMs) are particularly useful for developing a comprehensive understanding of how someone thinks, performs, and feels during an occupational task [18]. For instance, to understand MWL, MAMs combine several measures (physiological and subjective ratings, and performance), providing more reliable, valid, and comprehensive information than simply capturing and assessing data derived from a single source (e.g., subjective questionnaires) [2,15]. Multimodal data-fusion techniques can be helpful in fast-paced jobs, such as those in the military, healthcare, industrial technology, and public safety sectors, to sustain performance and mitigate critical errors [15]. Hence, integrating multiple types of measures enhances accuracy and reliability, thereby providing an all-encompassing picture of a person’s mental load in the workplace.

### 4.6. Artificial Intelligence & Machine Learning Methods

Data fusion techniques also enable the integration of ML techniques and AI methods, providing enhanced predictability for early warning signs and preemptive intervention for mental stress and other health-related issues, which is particularly useful for monitoring individuals in demanding occupations [2,6]. The fusion of multimodal data and methods used to integrate these data, such as ML and AI, can quickly show whether a person’s job performance drops a certain percentage below the established non-mental overload baseline (e.g., >20%), likely allowing for rapid early detection and strategic intervention to mitigate performance errors, sustain productivity, and ensure safety.

The use of data fusion techniques with ML and AI presents specific challenges that deserve consideration. Hence, to overcome potential misleading data output, it is important to

1. Maintain data integrity and continually update data across different locations and devices (data synchronization).
2. Ensure sensors are precisely calibrated and adjusted.
3. Workers performing the same occupational task can exhibit varying brain activities and stress response patterns [7,15].
4. Recognize how AI/ML systems may present false positives, introduce algorithmic bias, and are dependent on high-quality, large data sets representative of the population [23,38].

The future predictive capabilities of AI and ML techniques may become particularly relevant in applied workplace environments for real-time monitoring of cognitive stress, such as the following human-robot interaction case study.

### 4.7. Human-Robot Case Study: Tracking Cognitive Overload

This case study assessed the mental demands of a human interacting with a robotic arm, commonly referred to as a cobot, in an industrial smart factory. The case study explored probable methods for the objective measurement of mental stress of a human in real time [39].

In an industrial smart factory case study, a human operator (Lisa) coordinated with a cobot to complete time-sensitive classification and sorting tasks. While first appearing efficient, the setup overlooked a significant factor: the cobot operated at a fixed pace, disregarding the operator’s mental or physical state [39]. To evaluate RT mental load, the research team employed EEG and functional Near-Infrared Spectroscopy (fNIRS) technologies, alongside the inclusion of a foot pedal, to measure reaction time to audio stimuli [39].

This dual-task activity replicated actual factory work. For the primary task, the cobot handed Lisa color-coded boxes, and she had to quickly decide whether the printed equation on the box was correct. She then had to sort the box using either the text color or the color words written on it [39]. For the secondary task, she had to press a foot pedal below her factory workstation as quickly as possible to track her divided attention and stress level. If she fell behind by not matching the speed of the cobot, the boxes were dropped. The reaction time was over five episodes, each lasting four minutes [39]. The results of this case study depicted peak cognitive overload during rapid task sequences, suggesting the need for adaptive cobot behavior based on human stress signals [39].

Wearable sensors can also be integrated into other high-demand professions, such as healthcare, public safety, air traffic control, and biotechnology, to measure real-time cognitive loads. However, the sensor modalities used to measure cognitive load would vary based on job type. For example, a surgeon who performs complex surgeries for long periods of time does not wear an EEG cap during surgery and thus would require less invasive biosensor technologies to assess his cognitive load.

### 4.8. Adaptive Scheduling Using Real-Time Biometric Alerts

Adaptive scheduling uses RT biometric and neuroergonomic assessments to adjust tasks ahead of the user by measuring and predicting their combined workload. Insights from the revealed patterns inform the planning of challenging task demands and adverse working environments, including rest intervals, to mitigate stress and fatigue [8]. The advantages of RT alerting for supervisors and systems are extensive, because they can signal multiple task components across operations.

### 4.9. Continuous RT Monitoring & Performance

Cutting intermediaries between physiological data and synthesized results enables accurate biometric sensors to reduce costs and time and significantly preserve lives. In jobs that require intense physical or cognitive task execution, the continuous collection of RT data can provide a better understanding of the user’s current performance ability and potentially recognize red flag readings that indicate cognitive or physical fatigue [13]. In addition, the use of wearable biometric devices and improved employee onboarding processes have positively affected the industry. Complex and hazardous training scenarios can be broken down into more specific and digestible instructions through virtual simulations to strengthen familiarity with and experience with new workstations, operational processes, and equipment. This leads workers to achieve greater productivity more rapidly while reducing the time, money, and resources required [33].

### 4.10. Ethics and Data Governance

Addressing sensitive user data requires recognizing that a person’s health information is personal and confidential [21]. Many users are unaware of the privacy risks associated with wearable biometric devices and how their data are protected [9,22]. Biometric wearable device privacy policies are often difficult to read on small screens, which reduces user understanding and can lead to unfair targeting, misleading or persuasive advertising, and the potential loss of personal data [9,30]. Furthermore, different types of occupational ecosystems can pose challenges for biometric signaling wearables owing to overheating, water/sweat leakage, and a lack of sensor connectivity in jobs such as construction, healthcare, first responders, industrial tech factories, and mining [34], or in extreme environments such as space and deep-sea exploration.

Ethical implications arise when AI is used to process sensitive data. Integrating AI entails tracking data outside the necessary biometric measurements, particularly in surveillance and user behavior. AI-powered biometric monitoring can cause social harm when data from workers are kept from them and can be used to increase managerial control [3]. Employees should be able to review and consent to a predetermined list of trackable actions to ensure the proper use of AI systems and to avoid invasion of user health privacy. Further explorative studies are necessary to fully understand how AI monitoring may influence decisions made by industry executive leaders and employee beliefs. Preventive policy measures could help relieve uncertainties surrounding the opaque system’s purpose and ensure that employees do not have private health data used against them [3].

### 4.11. Future Directions

Personalizing mental load thresholds through individualized baseline modeling can help build consistent and accurate physiological profiles, while minimizing errors in generalized biometric systems [17]. This increases user trust in biometric outputs, even when occupational task demands vary. Hence, the integrative use of AI and ML can help create work schedules that match a person’s cognitive resources at a specific time, resulting in better task timing and reduced mental stress, while also aiding employee safety and allowing management leaders to make informed decisions based on RT data [2].

To develop a more holistic approach for predicting mental and physical performance workloads using AI and ML platforms, the fusion of various types of job-relevant data increases predictability and accuracy, thereby overcoming the limitations of relying solely on a unimodal approach [2]. Finally, differences in job roles and users imply that biometric variance is expected and must be accounted for [30].

## 5. Conclusion

This review examines the core theories supporting cognitive overload to better understand how task demands disrupt cognitive processing. By integrating multidimensional measures (physiological, behavioral, and subjective) and using AI and ML for multimodal data fusion, it is feasible to enhance the predictive accuracy of cognitive overload, thereby enabling prompt adaptive strategies in high-demand occupations. Although these advanced methods are promising, they also raise critical ethics and data governance concerns that must be addressed prior to large-scale deployment. Future longitudinal studies should prioritize real-world validation of these approaches, personalize individual cognitive workload thresholds, and establish transparent ethics and governance frameworks that safeguard private data while also supporting sustained cognitive and physical health.

## Data Availability

All data produced in the present work are contained in the manuscript.

## Appendix A. Oxygen measurements

**Table A1.**
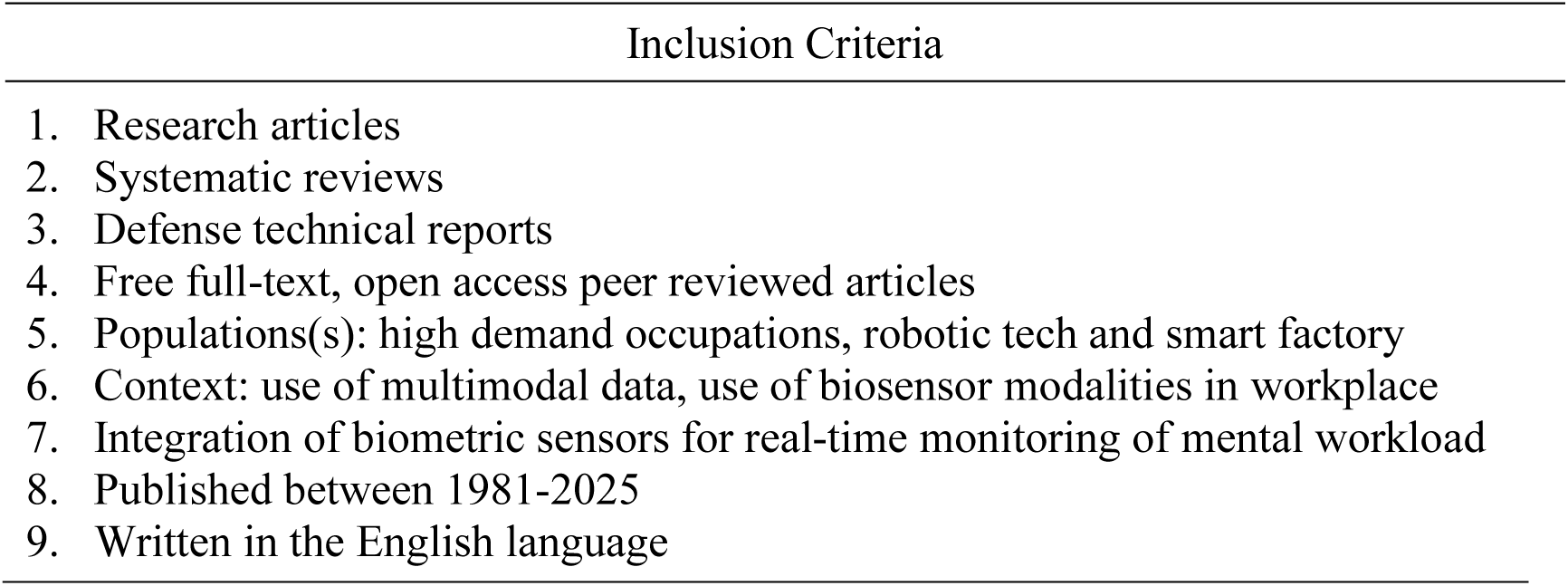
Inclusion Criteria.

| Inclusion Criteria |
| --- |
| 1. Research articles |
| 2. Systematic reviews |
| 3. Defense technical reports |
| 4. Free full-text, open access peer reviewed articles |
| 5. Populations(s): high demand occupations, robotic tech and smart factory |
| 6. Context: use of multimodal data, use of biosensor modalities in workplace |
| 7. Integration of biometric sensors for real-time monitoring of mental workload |
| 8. Published between 1981-2025 |
| 9. Written in the English language |

**Figure A1.**
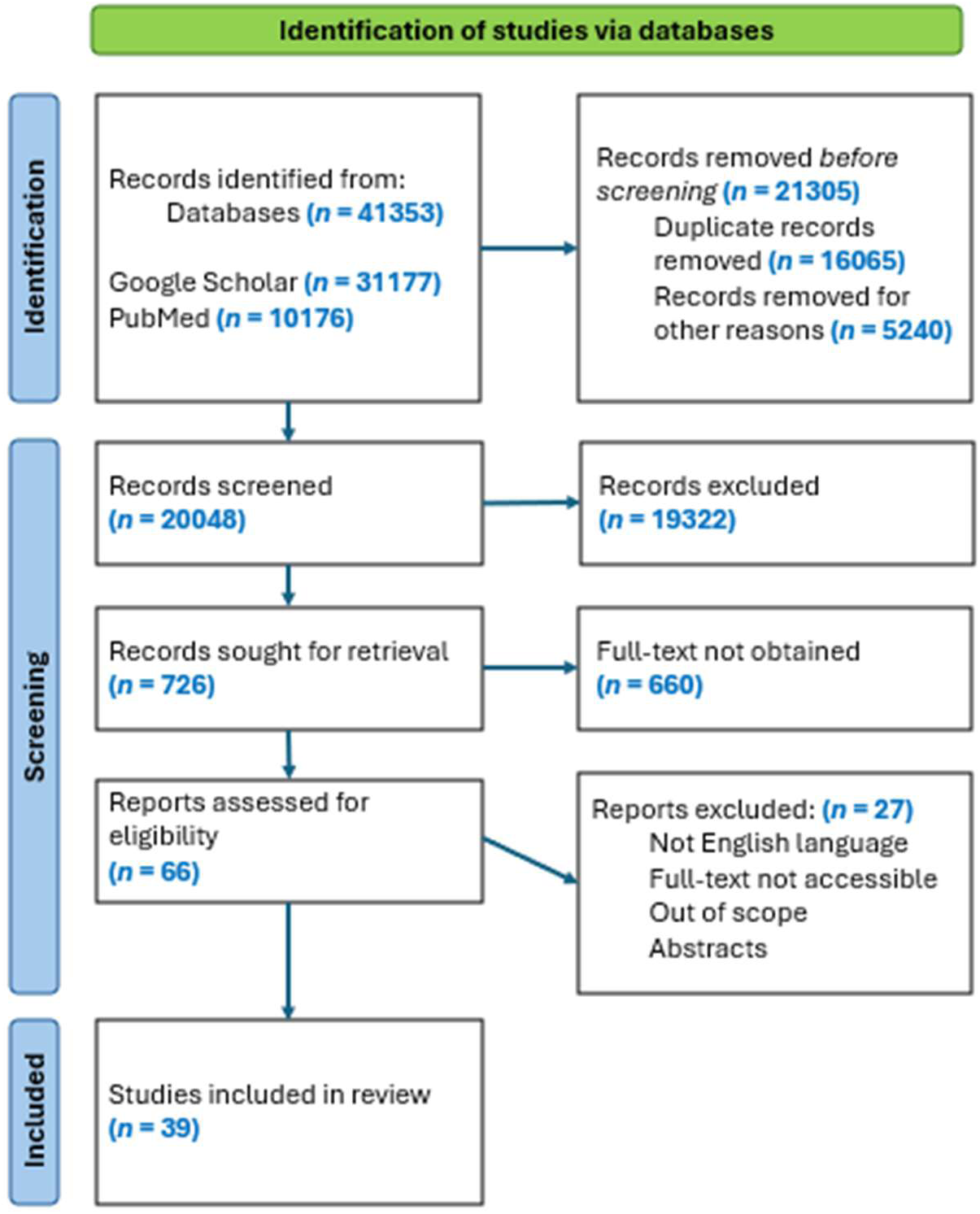
PRISMA Flow Diagram

